# Efficacy and safety of Ivermectin and Hydroxychloroquine in patients with severe COVID-19. A randomized controlled trial

**DOI:** 10.1101/2021.02.18.21252037

**Authors:** Jose Lenin Beltran Gonzalez, Mario González Gámez, Emanuel Antonio Mendoza Enciso, Ramiro Josue Esparza Maldonado, Daniel Hernández Palacios, Samuel Dueñas Campos, Itzel Ovalle Robles, Mariana Jocelyn Macías Guzmán, Andrea Lucia García Díaz, César Mauricio Gutiérrez Peña, Lucila Martinez Medina, Victor Antonio Monroy Colin, Arreola Guerra Jose Manuel

## Abstract

**Background:** In the search for active drugs against COVID-19, the indications of many have been redirected. Ivermectin and Hydroxychloroquine are drugs that inhibit viral replication *in vitro* and that have been used in several medical centers.

**Objectives:** This clinical trial analyzes the efficacy of Ivermectin and Hydroxychloroquine in patients with moderate COVID-19 and in need of hospitalization.

**Methods:** This a controlled, clinical, randomized, double-blind trial that included patients with COVID-19-induced pneumonia and hospitalization criteria, but no severe respiratory failure. Patients were randomized to one of three groups: Group1-hydroxychloroquine, 400 mg every 12 hours on the first day and subsequently, 200 mg every 12 hours for 4 days, Group 2-ivermectin, 12 mg or 18 mg, according to patient weight and, Group 3-placebo. At inclusion, blood samples for arterial blood gases and biochemical markers associated with a poor prognosis were obtained. The primary outcome was established as the duration of hospitalization until discharge due to patient improvement, the total duration of hospitalization, and the safety outcomes were either respiratory deterioration or death.

**Results:** During the month of August, the admission of patients requiring hospitalization mostly encompassed cases with severe respiratory failure, so we ended the recruitment process and analyzed the data that was available at the time. One hundred and six (106) patients with an average age of 53 yrs. (±16.9) were included, with a greater proportion of males (n=66, 62.2 %). Seventy-two percent (72%) (n= 76) had an associated comorbidity. Ninety percent (90 %) of patients were discharged due to improvement (n=96). The average duration of hospitalization was 6 days (IQR, 3 – 10). No difference in hospitalization duration was found between the treatment groups (Group1: 7 *vs* Group 2: 6 *vs* Group 3: 5, p=0.43) nor in respiratory deterioration or death (Group 1: 18 % *vs* Group 2: 22.2 % *vs* Group 3: 24.3 %, p =0.83).

**Conclusions:** In non-critical hospitalized patients with COVID-19 pneumonia, neither ivermectin nor hydroxychloroquine decreases the number of in-hospital days, respiratory deterioration, or deaths.

**ClinicalTrials identifier NCT04391127**

## Introduction

On January 30, 2020, the World Health Organization declared a global health emergency due to SARS-Cov-2 infections (COVID-19). (1) Since then, the outbreak has spread to all continents and the number of confirmed cases continues to increase. To date, there is no commercially available vaccine or approved treatment, and the management of patients that develop symptoms and require hospitalization is mostly supportive.

Chloroquine and hydroxychloroquine belong to the aminoquinoline drug family, and are broadly used as a result of their immunomodulatory and potentially antiviral effects and their well-established safety profile. Since the development of the SARS-CoV public health emergency in Southern China in 2002, these drugs were considered potentially therapeutic.(2) Chloroquine and its analog hydroxychloroquine have been proposed as a prophylactic and therapeutic alternative in the management of the different COVID-19 clinical presentations; however, they have not been shown to improve clinical outcomes.(3-5)

In the search for active drugs against COVID-19, the indications of several drugs have been redirected. One of the most studied is ivermectin, a macrolide obtained from *Streptomyces avermitilis*. The FDA (Food and Drug Administration) authorized the use of this antiparasitic agent in humans in 2008. Significant ivermectin *in vitro* antiviral effects against SARS-Cov-2 have been recently reported, leading to the development of several clinical studies to determine its efficacy in COVID-19. (6,7)

We aim to evaluate the efficacy and safety of ivermectin and hydroxychloroquine patients with pneumonia secondary to COVID-19.

## Methodology

This is a controlled, randomized, double-blind, clinical trial of patients with pneumonia secondary to SARS-CoV-2 infection and that fulfilled hospitalization criteria. These criteria were defined according to the attending physician in the emergency department and included the following parameters: severity of clinical presentation (determined with the CURB-65 scoring system), need for supplemental oxygen, the presence of comorbidities and, laboratory markers suggesting a poor prognosis.

The patients included in the study had to fulfill the operational definition of a suspected or confirmed COVID-19 case as well as the pneumonia ATS criteria.(1,8) The following patients were considered: 1) Positive RT-PCR for SARS-CoV-2 by nasal and oropharyngeal swabbing, 2) Pneumonia, diagnosed by X-ray or high-resolution chest CT scan, with a pattern suggesting involvement due to coronavirus, 3) Recently established hypoxemic respiratory failure or acute clinical deterioration of pre-existing lung or heart disease.

Patients were excluded if they required high oxygen volumes (face mask > 10 L/ min), if they had predictors of a poor response to high-flow oxygen nasal prong therapy or if they required mechanical ventilation.(9)

Patients were classified as high- or low-risk for the development of QT interval prolongation due to hydroxychloroquine, according to their electrocardiogram. The QT interval was measured with Bazett’s formula. Patients with an interval ≥ 500 ms were randomized to ivermectin or placebo, while those with an interval < 500 ms were randomized to ivermectin, hydroxychloroquine, or placebo. The dose of ivermectin was 12 mg in patients weighing less than 80 kg and 18 mg in those above 80 kg.(10)

In the hydroxychloroquine group, patients were administered 400 mg every 12 hours on the first day, followed by 200 mg every 12 hours for another 4 days. Due to the physical appearance of hydroxychloroquine, calcium citrate was chosen as a placebo and was administered as 2 tablets every 12 hours on the first day, followed by one tablet every 12 hours on the following 4 days. Blinding was assured with amber-colored vials. Each patient had a vial for the initial dose in order to blind the ivermectin and a second vial for subsequent doses. All patients received two vials, one with the initially prescribed dose and a second one, with the indication to take two tablets 12 hours after the initial dose followed by one tablet every 12 hours until all tablets were finished.

On admission, blood samples were obtained to determine arterial blood gases, a complete blood count, blood chemistry, and prognostic markers such as fibrinogen, D-dimer, ferritin, troponin I, procalcitonin, C reactive protein, prothrombin and activated partial thromboplastin times. If available, a high-resolution chest CT scan was also obtained and if not, only a chest X-ray. The diagnostic probability of pneumonia due to SARS-CoV-2 was established following the CO-RADS classification.(11)

All hospitalized patients received pharmacological thromboprophylaxis with low molecular weight heparin or unfractionated heparin according to local and international guidelines.(12,13)

During the last week of June and based on the RECOVERY trial, we initiated the administration of dexamethasone, 6 mg IV every 24 hours, for 10 days or until discharge, in patients requiring oxygen therapy.(14)

All included patients were under continuous cardiac monitoring and protocolized therapy was withdrawn in patients that developed any arrhythmia or acute coronary syndrome.

Safety outcomes were established as death due to any cause or respiratory deterioration. Respiratory deterioration was defined as a respiratory frequency above 30 breaths per minute, a required inspired oxygen fraction delivered by face mask or high-flow nasal prongs of 60% or above, a PaO2/Fio2 ratio <200, or a ROX index at 12 hours <3.85 points.(9,15)

Hospital discharge was considered when the patient fulfilled the following criteria: absence of neurologic complications, no fever, hemodynamic stability over at least, the previous 72 hours, minimal oxygen requirements (nasal prongs at 1–2 liters per minute), and the availability of a well-established social support network.

The main outcome was determined as the hospitalization duration until discharge due to clinical improvement, the total duration of hospitalization, and the safety outcomes were duration of hospitalization until respiratory deterioration (previously defined) or death. This study was conducted at the *Hospital Centenario Miguel Hidalgo* in the state of Aguascalientes (Mexico), a tertiary care institution for the population lacking social security.

The study protocol was approved by the Ethics Committee of the *Hospital Centenario Miguel Hidalgo* on April 15, 2020, with the assigned number 2020-R-24. It was also included in the *ClinicalTrials* website with the identifier NCT04391127.

### Statistical Analysis

Depending on the measurement level, descriptive statistics were used. The distribution of continuous variables was determined with the Kolmogorov Smirnov test. Continuous variables with a normal distribution are expressed in means and their standard deviation while those with an abnormal distribution, as medians and their interquartile ranges. Categorical variables are presented as relative and absolute frequencies. Between-group analysis was evaluated with variance analysis (ANOVA) or the Kruskal Wallis test, depending on the distribution. Dichotomous or ordinal variables were analyzed by chi-square or Fisher’s exact test, as needed. Survival analysis was performed for the outcomes of death or respiratory deterioration, with Kaplan Meier curves, and between-group comparisons were obtained with the Log-rank test. A p value below 0.05 was considered significant. Microsoft Excel 2013 and STATA version 11.1 software were used for analysis.

Considering a mean hospitalization stay of 20 to 30 days and standard deviation of 7 days, if we want a reduction of 4 days of hospitalization, taking into account the mean differences formula, we calculate 47 patients per group of treatment.

## Results

During the past month of August, we observed a very significant decrease in the number of potential candidates that could be included in the study, since practically all hospital admissions required therapy with high oxygen concentrations or invasive mechanical ventilation. Based on the Ethics Committee’s recommendations, we decided to end recruitment and conduct an analysis with the data obtained as of August 15, 2020.

At the time of analysis, 108 patients had been recruited, two of which were eliminated because they were transferred to another hospital. The patients’ average age was 53 years (±16.9), and there was a greater proportion of males (n=66, 62.2 %). Comorbidities were present in 72% of cases (n= 76). Type 2 diabetes mellitus and systemic arterial hypertension were the most frequent (33.9 and 32.1 %). Mean body weight was 82.3 kg (±19.6) with a BMI of 29.6. (±6.6) (Table1).

**Table 1.** **Population characteristics. SAH: Systemic Arterial Hypertension, CKD: Chronic Kidney Disease, COPD: Chronic Obstructive Pulmonary Disease, BMI: Body Mass Index**

**Figure 1. Time to compound outcome of death or respiratory deterioration (p=0.44)**
| Variables | Entire group<br>(n= 106) | Hydroxychloroquine<br>(n=33) | Ivermectin<br>(n= 36) | Placebo<br>(n= 37) | P |
| --- | --- | --- | --- | --- | --- |
| Age, m (±SD) | 53.8 (16.9) | 48.9 (15.3) | 56 (16.5) | 53.8 (16.9) | 0.15 |
| Males, n (%) | 66 (62.2) | 22 (66.6) | 21 (58.3) | 23 (62.1) | 0.77 |
| Diabetes Mellitus, n (%) | 36 (33.9) | 11 33.3) | 9 (25) | 16 (43.2) | 0.25 |
| SAH, n (%) | 34 (32.1) | 8 (24.2) | 12 (33.3) | 14 (37.8) | 0.47 |
| CKD, n (%) | 5 (4.7) | 2 (6.1) | 2 (5.5) | 1 (2.7) | 0.73 |
| COPD, n (%) | 7 (6.6) | 1 (3) | 2 (5.5) | 4 (10.8) | 0.501 |
| Healthy, n (%) | 30 (28.3) | 13 (39.3) | 10 (27.7) | 7 (18.9) | 0.17 |
| Weight, m (±DE) | 82.3(19.6) | 84.8 (20.8) | 80 (19.7) | 82 (18.2) | 0.66 |
| BMI, m (±DE) | 29.6 (6.6) | 30.3 (6.3) | 29.2 (7) | 29.4 (6.6) | 0.55 |

A high-resolution chest CT scan was obtained in 93 patients. On admission, 69 patients had typical infiltrates characteristic of SARS-CoV-2 infection pneumonia, defined by CO-RADS 5, while 24 patients had an image with a low probability of SARS-CoV-2 infection according to that scoring system, but they had a diagnostically compatible clinical presentation and/or a positive RT-PCR test for SARS-CoV-2. (Table 2)

**Table 2.** Computed Tomography Characteristics.

| **Variables** | **Entire group (n= 93)** | **Hydroxychloroquine (n=28)** | **Ivermectin (n= 30)** | **Placebo (n= 35)** |
| --- | --- | --- | --- | --- |
| **CORADS 2, n (%)** | 5 (5.3) | 0 | 3 (10) | 2 (5.2) |
| **CORADS 3, n (%)** | 15 (16.1) | 4 (14.2) | 7 (23.3) | 4 (11.4) |
| **CORADS 4, n (%)** | 19 (20.4) | 7 (25) | 4 (13.3) | 8 (22.8) |
| **CORADS 5, n (%)** | 50 (53.7) | 17 (25) | 13 (43.3) | 20 (57.1) |
| **CORADS 6, n (%)** | 4 (4.3) | 0 | 3 (10) | 1 (2.8) |

The ratio between the Arterial Oxygen pressure (PaO2) and the Inspired Oxygen fraction (PaO^2^/FiO^2^) was greater than 200 mmHg in 64 patients; 42 patients had severe respiratory deterioration (Table 3).

**Table 3.** **Biochemical and gasometric markers. O2Sat: Oxygen saturation, PaO2/FiO2: Index of Oxygen arterial pressure / inspired oxygen fraction, PCO2: Carbon dioxide pressure, Hb: Hemoglobin, LDH: Lactic Dehydrogenase, CRP: C reactive protein**

| Variables | Entire group<br>(n= 106) | Hydroxychloroquine<br>(n=33) | Ivermectin<br>(n= 36) | Placebo<br>(n= 37) | P |
| --- | --- | --- | --- | --- | --- |
| SatO2, m (±SD) | 84 (7) | 86 (9) | 83 (8) | 83 (8) | 0.27 |
| PaO2/FiO2, m (±DE) | 223 (103) | 224 (116) | 245 (107) | 201 (82) | 0.17 |
| <100, n (%) | 11 (10.3) | 5 (15.1) | 2 (5.5) | 4 (10.8) | 0.44 |
| 100 – 200, n (%) | 31 (29.2) | 8 (24.2) | 9 (25) | 14 (37.8) | 0.36 |
| >200 – 300, n (%) | 48 (45.2) | 17 (51.5) | 16 (44.4) | 15 (40.5) | 0.64 |
| > 300, n (%) | 16 (15.1) | 3 (9.1) | 9 (25) | 4 (10.8) | 0.14 |
| PCO2, m (±SD) | 29 (7.5) | 26.3 (7.9) | 30.4 (7.2) | 30.3 (6.8) | 0.09 |
| Lactate, m (±SD) | 1.41 (0.6) | 1.45 (0.7) | 1.2 (0.6) | 1.5 (0.6) | 0.31 |
| Hb, m (±SD) | 13.5 (2.9) | 13.6 (3) | 13.1 (3) | 13.7 (2.8) | 0.63 |
| Leukocytes, m (±SD) | 10 (4.4) | 9.6 (3.5) | 10.3(5.1) | 10.1 (4.5) | 0.92 |
| Neutrophils, m (±SD) | 8 (4.1) | 7.7 (3.4) | 8.4 (4.8) | 7.8 (4.1) | 0.82 |
| Lymphocytes, m (±SD) | 1.3 (0.6) | 1.3 (0.54) | 1.2 (0.6) | 1.4 (0.7) | 0.40 |
| Platelets, m (±SD) | 254 (104) | 250 (91) | 261 (100) | 251 (119) | 0.90 |
| Creatinine, m (±SD) | 1.48 (2.8) | 0.95 (0.7) | 1.6 (3.3) | 1.8 (3.4) | 0.81 |
| LDH, m (±SD) | 394 (158) | 394 (189) | 369 (123) | 394 (158) | 0.19 |
| D-Dimer, m (±SD) | 1618 (1442) | 1593 (1845) | 1872 (1137) | 1380 (1268) | 0.01 |
| Fibrinogen, m (±SD) | 441 (247) | 439 (249) | 473 (250) | 413 (244) | 0.53 |
| **CRP, m (±SD)** | 117 (136) | 171 (122) | 187 (126) | 172 (158) | 0.67 |
| **Ferritin, m (±SD)** | 721 (927) | 669 (667) | 902 (1125) | 592 (910) | 0.35 |
| **D Bilirubin, m (±SD)** | 0.64 (1.6) | 0.91 (2.9) | 0.53 (0.5) | 0.5 (0.6) | 0.56 |
| **Troponin, m (±SD)** | 0.05 (0.2) | 0.1 (0.3) | 0.03 (0.08) | 0.02 (0.02) | 0.18 |

Patients included in the study had characteristics considered significantly severe: in 77%, the Neutrophil / Lymphocyte index was ≥ 3, SOFA ≥ 2 in 93 %, APACHE ≥ 8 points in 96 % and CURB-65 ≥2 points in 34.9 %. The score in severity scales was not different between-groups (Table 4)

**Table 4.** Markers and Prognostic Score Systems.

| **Variables** | **Entire**<br>**group (n=**<br>**106)** | **Hydroxychloro**<br>**quine**<br>**(n=33)** | **Ivermectin**<br>**(n= 36)** | **Placebo**<br>**(n= 37)** | **P** |
| --- | --- | --- | --- | --- | --- |
| **N/L Index ≥ 3, n (%)** | 89 (77.7) | 30 (90.9) | 31 (86.1) | 28 (77.7) | 0.34 |
| **SOFA, m (SD)** | 3.3 (1.6) | 3.1 (1.5) | 2.9 (1.5) | 3.8 (1.7) | 0.05 |
| **SOFA < 2, n (%)** | 7 (6.6) | 2 (6.1) | 4 (11.1) | 1(2.7) | 0.33 |
| **2 – 3, n (%)** | 61 (57.6) | 21 (63.6) | 22 (61.1) | 18 (48.6) | 0.38 |
| **≥ 4, n (%)** | 38 (35.8) | 10 (30.3) | 10 (27.7) | 18 (48.6) | 0.14 |
| **APACHE II, m8 -15**<br>**(±SD), n (%)** | 11.2 (4)<br>67 (63.2) | 11.1 (3.8)<br>22 (66.6) | 10.6 (3.7)<br>23 (63.8) | 11.9 (4.4)<br>22 (59.4) | 0.38<br>0.14 |
| **>15, n (%)** | 23 (21.2) | 7 (21.2) | 5 (13.8) | 11 (21.7) | 0.25 |
| **CURB65, m (±SD)** | 1.08 (1.03) | 0.9 (0.9) | 1.1 (1) | 1.1 (1.1) | 0.77 |
| **≥2, n (%)** | 37 (34.9) | 11 (33.3) | 14 (38.8) | 12 (32.4) | 0.82 |

During hospitalization, 52.3% of patients received some form of antibiotic, 92.2% were on thromboprophylaxis and 55.7% received dexamethasone as ancillary therapy (Table 5).

**Table 5.**
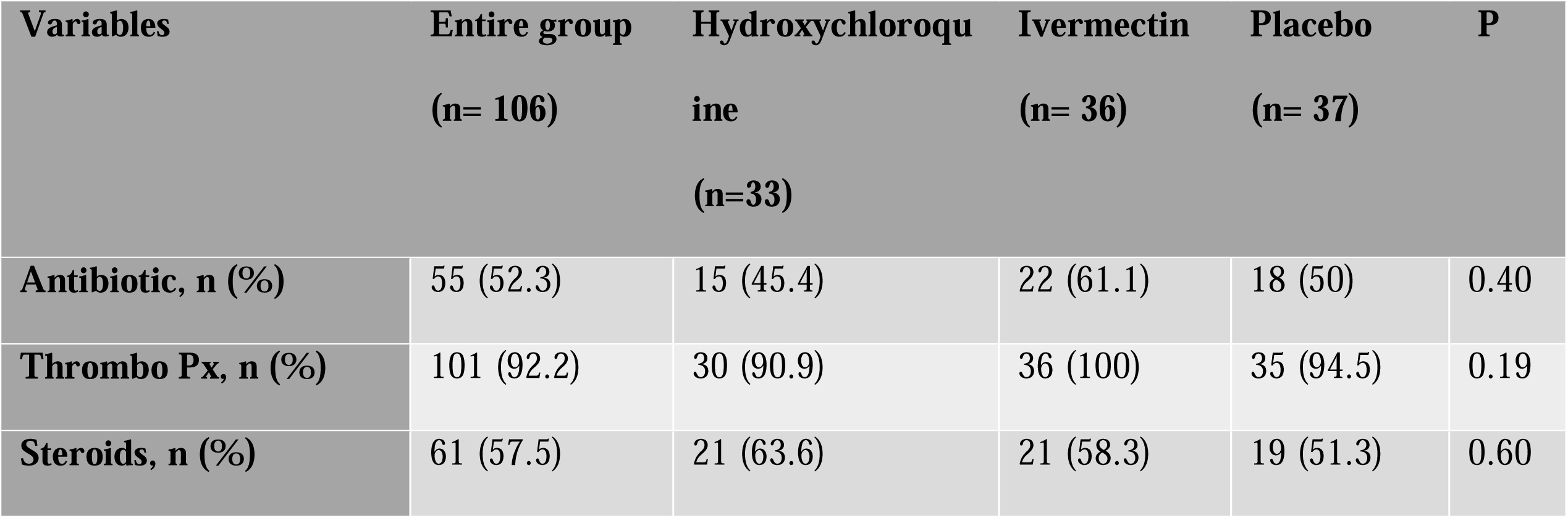
Ancillary Treatments.

| **Variables** | **Entire group**<br>**(n= 106)** | **Hydroxychloroquine**<br>**(n=33)** | **Ivermectin**<br>**(n= 36)** | **Placebo**<br>**(n= 37)** | **P** |
| --- | --- | --- | --- | --- | --- |
| **Antibiotic, n (%)** | 55 (52.3) | 15 (45.4) | 22 (61.1) | 18 (50) | 0.40 |
| **Thrombo Px, n (%)** | 101 (92.2) | 30 (90.9) | 36 (100) | 35 (94.5) | 0.19 |
| **Steroids, n (%)** | 61 (57.5) | 21 (63.6) | 21 (58.3) | 19 (51.3) | 0.60 |

Follow-up during hospitalization did not reveal thrombotic complications and no arrhythmias developed in patients on hydroxychloroquine. Only 3 patients developed septic shock, one in each treatment arm.

### Outcomes

The average duration of hospitalization was 6 days (IQR 3 – 10). Ninety percent (90 %) of patients were discharged after clinical improvement (n=96). Twenty-three (23) patients developed respiratory deterioration, 13 (12.2%) of whom died during follow-up, all as a result of respiratory failure, and three with sepsis due to bacterial coinfection. No differences in outcome were detected between the treatment groups (Table 6). The time until death or respiratory deterioration was also not different between groups (figure 1).

**Table 6.** Outcomes.

| **Outcome** | **Entire group (n= 106)** | **Hydroxychloroquine (n=33)** | **Ivermectin (n= 36)** | **Placebo (n= 37)** | **p value** |
| --- | --- | --- | --- | --- | --- |
| **Duration of Hospitalization, med (IQR)** | 6 (3 – 10) | 7 (3 – 9) | 6 (4 – 11) | 5 (4 – 7) | 0.43 |
| **Hospital discharge, n (%)** | 96 (90.5) | 30 (90.9) | 32 (88.8) | 34 (91.8) | 0.91 |
| **Discharge without respiratory deterioration or death, n (%)** | 80 (75.4) | 26 (78.7) | 27 (75) | 27 (72.9) | 0.85 |
| **Respiratory deterioration or death, n (%)** | 23 (21.7) | 6 (18.1) | 8 (22.2) | 9 (24.3) | 0.83 |
| **Death, n (%)** | 13 (12.2) | 2 (6) | 5 (13.8) | 6 (16.2) | 0.42 |

**Figure 1.**
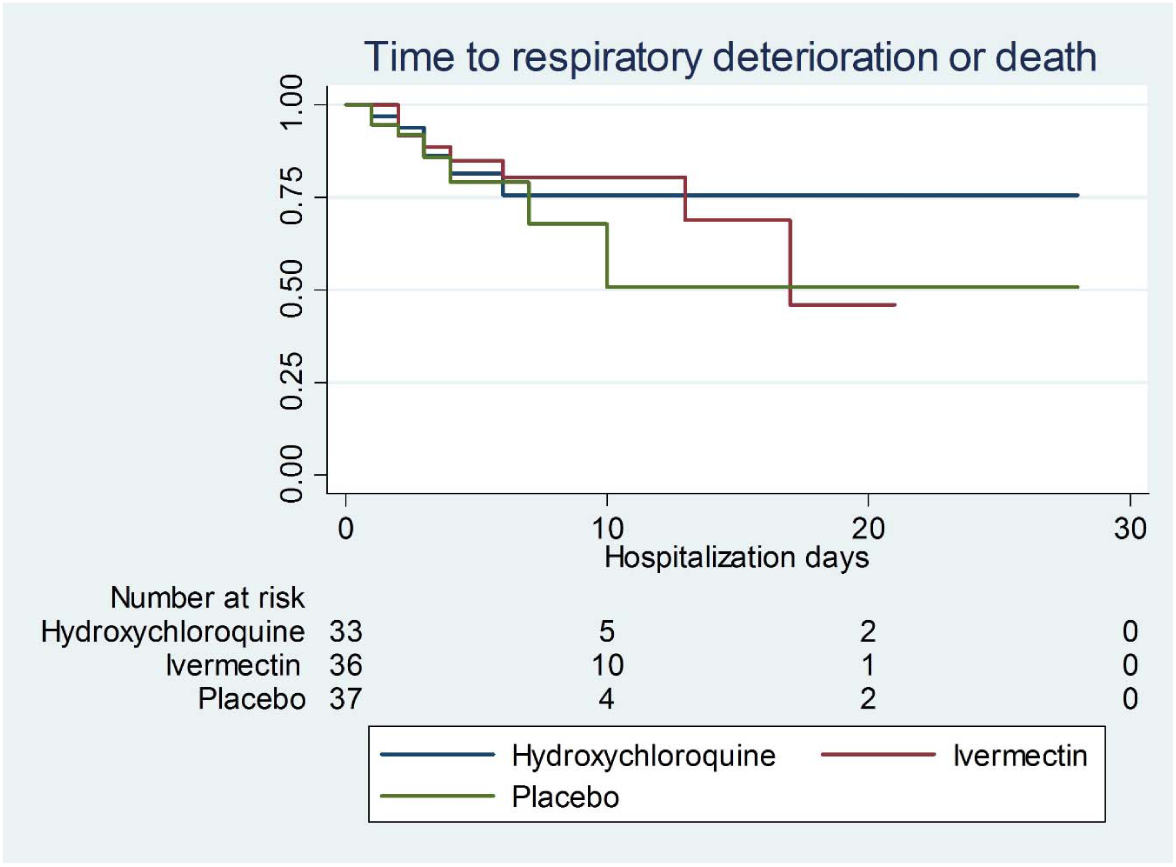
Time to compound outcome of death or respiratory deterioration (p=0.44)

## Discussion

In this study of non-critically ill patients with pneumonia secondary to COVID-19 and fulfilling hospitalization criteria, treatment with hydroxychloroquine or ivermectin was not superior to placebo, neither in terms of hospitalization duration nor in progression to severe respiratory failure or death.

In the first weeks of the COVID-19 pandemic, there was *in vitro* evidence on the efficacy of hydroxychloroquine as well as case series and non-controlled comparative studies supporting its use. This led various medical centers and health systems to recommend it based on compassionate use. This strategy fostered panic purchases and prescriptions leading to drug shortages for patients with well-established indications. As months went by, its inefficacy was suspected and reports from the SOLIDARITY study finally proved its therapeutic futility in decreasing mortality, and patient recruitment was stopped.(16) Our analysis further confirms that study’s findings.(4,5)

Although adverse events have been reported with the use of hydroxychloroquine, particularly QT interval prolongation, none of our patients developed cardiovascular complications associated with hydroxychloroquine use; perhaps, electrocardiographic screening may have contributed to the avoidance of these complications.

As with hydroxychloroquine, ivermectin was proposed in the early phases of the pandemic, as treatment and even prophylaxis of SARS-CoV-2 infection. It has mainly been used in Latin America as compassionate treatment with no evidence supporting its efficacy in COVID-19; some health ministries have even modified their treatment policies recommending its generalized use.(17)

As in the case of hydroxychloroquine, evidence in favor of ivermectin use was the result of *in vitro* studies and non-controlled comparative trials.(18) A possible explanation for those findings is the fact that the *in vitro* efficacy of ivermectin in decreasing the viral load of SARS-CoV-2, is clinically translated in much greater ivermectin doses *in vivo*, an unplausible proposal in the clinical setting.(19) To our knowledge, our study is the first clinical trial comparing ivermectin with placebo.(20)

The characteristics of patients enrolled in our study differ from those in series published in China and some European countries early on since our population has a greater incidence of overweight and obesity.

They have a greater load of comorbid disease thus impacting clinical results. As in the initial case series, systemic arterial hypertension and type 2 diabetes mellitus were the main entities compromising the patients’ course and fostering their development of respiratory insufficiency and an increase in adverse clinical outcomes.

The main strength of this clinical trial is the fact that it represents the response of a referral hospital in the management of patients with COVID-19. In a controlled manner, patients were offered two therapeutic alternatives that at the beginning of the pandemic appeared to be potentially effective. Although the patient number is not sufficient to reach categorical conclusions, the study’s design certainly suggests that both drugs are ineffective. We hope it will contribute to meta-analyses that may yield more robust conclusions.

The study’s main weakness is the limited number of patients per group; also, among the pre-established outcomes, we were unable to determine whether the SARS-CoV-2 PCR tests became negative, due to the lack of reactants and the minimal usefulness of proving its negativity from a clinical-practical viewpoint.

In conclusion, in non-critically ill, hospitalized patients with pneumonia secondary to COVID-19, the use of hydroxychloroquine or ivermectin did not decrease the number of hospitalization days, respiratory deterioration, or deaths.

## Data Availability

The date that could be shared with exception of patients name or file number. the data can be requested directly to the email of the corresponding author

